# Deep Learning based Quantification of Root Exposure in Lower Anterior Teeth using Intraoral camera image

**DOI:** 10.64898/2026.07.02.26357104

**Authors:** Hyunsuk Choi, Youn-Hee Choi, Eun Young Park, Sohee Kang, Eun-Kyong Kim

## Abstract

This study compared and evaluated two widely used deep learning-based artificial intelligence (AI) models, U-Net++ and YOLOv11, for quantifying tooth and root exposure on intraoral camera images of the mandibular anterior lingual region. Intraoral images of the mandibular anterior lingual region were collected from 291 patients (mean age, 52.8 years) at a university hospital dental clinic with institutional review board approval (YUMC IRB 2021-07-019-002). A total of 266 eligible images (mean, 5.50 teeth per image; 3.70 teeth with root exposure) were annotated. YOLOv11 and U-Net++ were fine-tuned using five-fold cross-validation with data augmentation. Model performance was evaluated on a held-out test set of 40 images using Dice coefficient, Intersection over Union (IoU), accuracy, mean Average Precision at an IoU threshold of 0.5 (mAP50), Lin’s concordance correlation coefficient (CCC), and intraclass correlation coefficient (ICC). Confidence intervals were estimated using 10,000 bootstrap iterations. For tooth segmentation, U-Net++ demonstrated superior performance, with high accuracy (0.981), Dice coefficient (0.971), and IoU (0.944). In contrast, for root segmentation, YOLOv11 outperformed U-Net++, achieving higher Dice (0.860 vs. 0.746) and IoU (0.762 vs. 0.631). Notably, YOLOv11 showed stronger agreement with the ground truth for quantifying the exposed root ratio (ERR) (CCC, 0.973; ICC, 0.975). These findings suggest that accurate detection of root exposure is important for assessing periodontal tissue loss and that YOLOv11 is a promising model for root exposure quantification in intraoral images. YOLOv11-based quantification of root exposure may serve as a useful adjunctive AI tool for screening and monitoring periodontal conditions and may support individualized treatment planning.

## 1. Introduction

Mandibular anterior root exposure, resulting from gingival recession, is a prevalent clinical condition primarily driven by chronic periodontitis, excessive tooth brushing, or orthodontic treatment[1,2]. This phenomenon leads to aesthetic concerns, dentin hypersensitivity, increased risk of root caries, and challenges in maintaining oral hygiene[3–5]. The exposed root surfaces exacerbate periodontitis through a bidirectional effect, with factors such as root concavities or plaque deposition leading to calculus formation contributing to disease progression[6]. Notably, lingual root exposure in the mandibular anterior region is reported to be significantly associated with chronic periodontitis rather than mechanical abrasion factors like excessive brushing, whereas buccal root exposure correlates more with dental wear[7–9]. Thus, detecting lingual root exposure in this region might be clinically significant for assessing the severity and progression of chronic periodontitis.

Traditionally, periodontal disease is diagnosed by probing the periodontal pocket or taking dental x-ray by dentist. By leveraging cone-beam computed tomography (CBCT) or radiographic imaging, AI can be used as an aid in measuring alveolar bone thickness, root distance, and exposure ratios, facilitating precise treatment planning, predicting tooth movement rates, and enhancing implant success rates[10–13]. However, visiting the dental hospital and taking an x-ray are not easy for the general public, so they tend to be reluctant and miss the right time for treatment. Therefore, quantifying mandibular anterior root exposure in home might offer substantial clinical benefits by motivating people. Specifically quantification using oral camera image and artificial intelligence (AI) models might support minimally invasive techniques, reduce patient morbidity, and promote evidence-based screening[14,15].

So far, AI has been widely applied in periodontal diagnostics, including alveolar bone loss detection on radiographs, with promising diagnostic performance reported across studies. AI’s contributions to tooth prognosis and bone density analysis further suggest significant potential for future research in this area [11,16–21]. Recently, we reported that deep learning can be used to quantify exposed root area from intraoral photographs of the mandibular anterior region and that the resulting exposed root ratio (ERR) serves as a useful biomarker for periodontitis risk prediction when combined with age [22]. However, that study relied on a single segmentation model (YOLOv11) without comparing alternative deep learning architectures. Because the choice of segmentation paradigm can substantially influence the accuracy and reproducibility of root exposure quantification, it remains unclear which approach is most suitable for this specific task.

In medical imaging, YOLO models excel in rapid object detection, while U-Net++ provides superior accuracy in medical image segmentation[23–25]. In dental imaging, YOLO is applied for detecting conditions like caries and periodontal bone loss[18,26–28], whereas U-Net++ is used in precise segmentation tasks, such as tooth and lesion delineation[17,29–31]. Considering that both models are frequently used for medical image analysis in the dental field, a comparison between the two models would be necessary.

Therefore, the present study aimed to compare the performance of two widely used deep learning models—YOLOv11 (instance segmentation) and U-Net++ (semantic segmentation)—for tooth and root exposure segmentation on intraoral camera images, with particular emphasis on the reliability of the derived ERR.

## 2. Methods

### 2.1. Study Participants

This study recruited a total of 315 patients aged over 35 years who visited the dental clinic of a university hospital between December 2021 and December 2022. Participants were enrolled as a convenience series, and the intraoral images were collected prospectively and analyzed retrospectively. No formal a priori sample-size calculation was performed; all eligible images acquired during the study period were included, and five-fold cross-validation was used to make the fullest use of the limited dataset. Of these, 291 patients (65 males and 226 females; mean age ± standard deviation [SD] = 52.8 ± 17.32 years) were included for the development of the deep learning models. Participants were included if they had at least two mandibular anterior teeth, no infectious diseases, and did not present with excessive calculus that obscured the exposed root surface identification. For generalization in clinical field, a little calculus was allowed in case of CEJ (cementoenamel junction) is visible. Written informed consent was obtained from all participants after explaining the purpose and methods of the study. Before image acquisition, each participant performed a water rinse to remove food debris. Subsequently, intraoral images focusing on the lingual surfaces of the mandibular incisors were captured using a professional intra-oral camera at a resolution of 1280 × 720 pixels.

The study protocol received approval from the institutional review board of Yeungnam University Hospital (approval no. YUMC IRB 2021-07-019-002). This diagnostic/prognostic investigation was performed in compliance with the ethical principles outlined in the Declaration of Helsinki [32] and adhered to the Standards for Reporting of Diagnostic Accuracy Studies (STARD) guidelines [33].

### 2.2. Dataset and annotation

Finally, images did not have at least one tooth with exposed root surface were excluded from dataset. Excluding images without exposed root structures from the training dataset may offer advantages in terms of improving root detection performance, enhancing loss function stability, and increasing training efficiency. Therefore, the dataset comprised 266 intraoral images (25 of the 291 patients were excluded because no tooth exhibited an exposed root surface), which were randomly divided into training and test sets with 226, 40 images ahead of model training. For annotation, a single experienced dentist conducted instance segmentation for all images, precisely delineating tooth and exposed root surfaces. This ground-truth annotation was performed independently of, and prior to, any model prediction. An open-source annotation platform, the Image Annotator (Bhattiprolu, 2024), was employed for this task.

### 2.3. Development of deep learning models

Two popular convolutional models were fine-tuned to segment teeth and exposed root surfaces on intraoral camera image dataset except held-out test set: (i) an Ultralytics YOLO-v11 model for object-wise segmentation, and (ii) a U-Net++ model for pixel-wise semantic segmentation. Both models were fine-tuned using five stratified splits (80/20 train/validation per split) constructed with 5-fold cross validation algorithm considering the distribution of Tooth/Root across folds.

First, for the YOLOv11 segmentation model, weights pre-trained on the MS COCO dataset were adopted for the backbone and neck using the ‘YOLO11m-seg.pt’ weight file downloaded from the Ultralytics GitHub repository (Jocher & Qiu, 2024). Training process adopted 768 as image size, early stopping, warm-up epochs, 0.001 as initial learning rate, decayed to 1% via a cosine schedule, momentum 0.937, and weight decay 5×10⁻⁴. On-the-fly augmentations included rotations (±15°), scaling (±10%), shearing, horizontal/vertical flips, and brightness modulation (HSV-V) [34]. After training using each split, the best weight file was selected by validation performance.

Second, U-Net++ with a ResNet-34 encoder pre-trained on ImageNet [35] was fine-tuned to predict three logits (background/Tooth/Root). Images were resized to 768 and padded to square; normalization followed ImageNet statistics. Training employed AdamW [36] with differential learning rates (5×10⁻⁵ for encoder; 1×10⁻⁴ for decoder/segmentation head) and weight decay 1×10⁻⁴. A cosine-annealing schedule with warm restarts was used initially, then switched to cosine annealing for the remaining epochs; early stopping monitored mean IoU of Tooth and Root with patience = 20. Augmentations were also applied (rotation ±15°, scale ±10%, shearing, horizontal/vertical flips, random brightness), with padding to maintain 768×768 input.

Each model was developed using Google Colab (Google LLC, Mountain View, CA, USA), a cloud-based platform for AI research. The hardware specifications of this environment included an NVIDIA A100-SXM4-40GB GPU (graphics processing unit) and an Intel(R) Xeon(R) CPU (central processing unit). The software specifications included with Python 3.12.11, torch 2.8.0+cu126, segmentation-models-pytorch 0.5.0, Ultralytics version 8.3.183, and CUDA 12.4. Python programming language (Python Software Foundation, Beaverton, OR, USA) were used for model development.

### 2.4. Comparison of model performance

To compare the performance of the fine-tuned YOLOv11 and U-Net++ models, we generated ground-truth masks for the “Tooth” and “Root” classes from the held-out test set annotations. Then we compared these masks with those predicted by each model at both pixel and instance level, respectively. Key metrics including Dice coefficient, Intersection over Union (IoU), pixel accuracy, and specificity were calculated [37]. Also, for instance-based metrics, mAP50 was calculated by evaluating predicted segmentation masks against ground truth masks using an IoU threshold of 0.5. Average Precision was computed from the precision-recall curve for each class, and mAP50 was the average of these class APs.

### 2.5. Quantification of Exposed Root Ratio (ERR)

To enable quantitative assessment of root exposure in dental imaging, the predicted areas of the tooth and exposed root were calculated for each model using the held-out test set. Based on these results, the ERR was calculated for ach image. For validation, the Intraclass Correlation Coefficient (ICC) and the Concordance Correlation Coefficient (CCC) [38,39] were computed against the ground truth values to evaluate both the reliability and agreement of the model outputs.

### 2.6. Statistical analyses

The bootstrap resampling method [40] was applied to estimate performance indicators and to calculate the ICC and CCC. A total of 1,000 bootstrap resampling iterations were used to estimate the performance indicators, while 10,000 iterations were performed to compute the ICC and the CCC. This extensive resampling procedure ensured stable estimates and reliable confidence intervals, although not used as a formal test.

## 3. Results

### 3.1. Dataset characteristics

The flow of patient and image selection is shown in Fig 1. Across the entire dataset, the mean number of teeth per image was 5.50 (SD = 0.79), with no difference observed between the training/validation and test subsets (Table 1). On average, 3.70 teeth (SD = 1.31) per image exhibited root exposure, with similar distributions in both the training/validation set (3.69 ± 1.31) and the test set (3.73 ± 1.32). These results indicate that the dataset was balanced across subsets with respect to tooth count and the prevalence of exposed roots, ensuring comparability in subsequent model evaluations.

**Fig 1.**
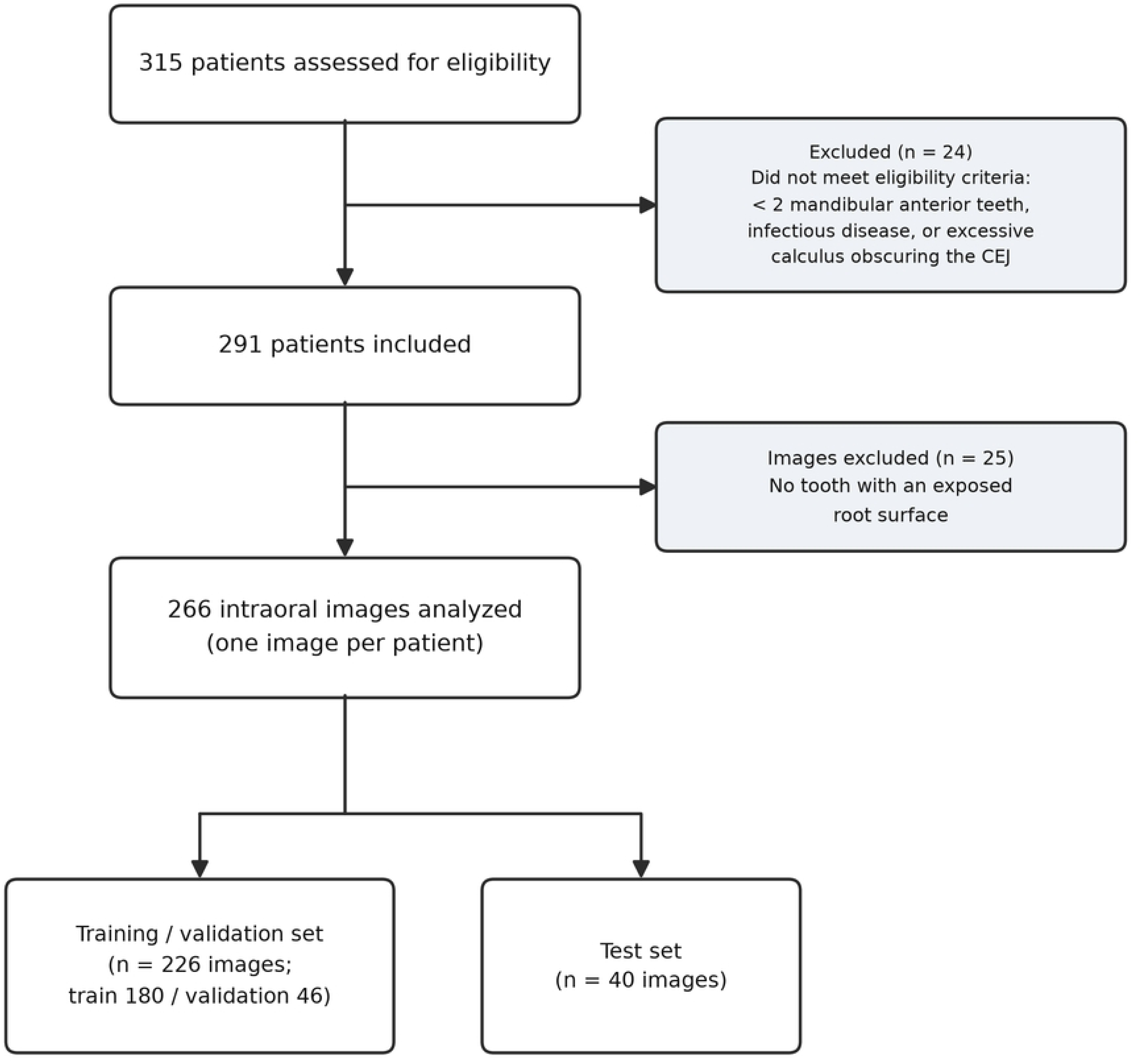
Flow diagram of patient and image selection (STARD). A total of 315 patients were assessed for eligibility and 291 were included; 266 intraoral images (one per patient) were then analyzed and divided into training/validation (n = 226) and test (n = 40) sets.

**Table 1.**
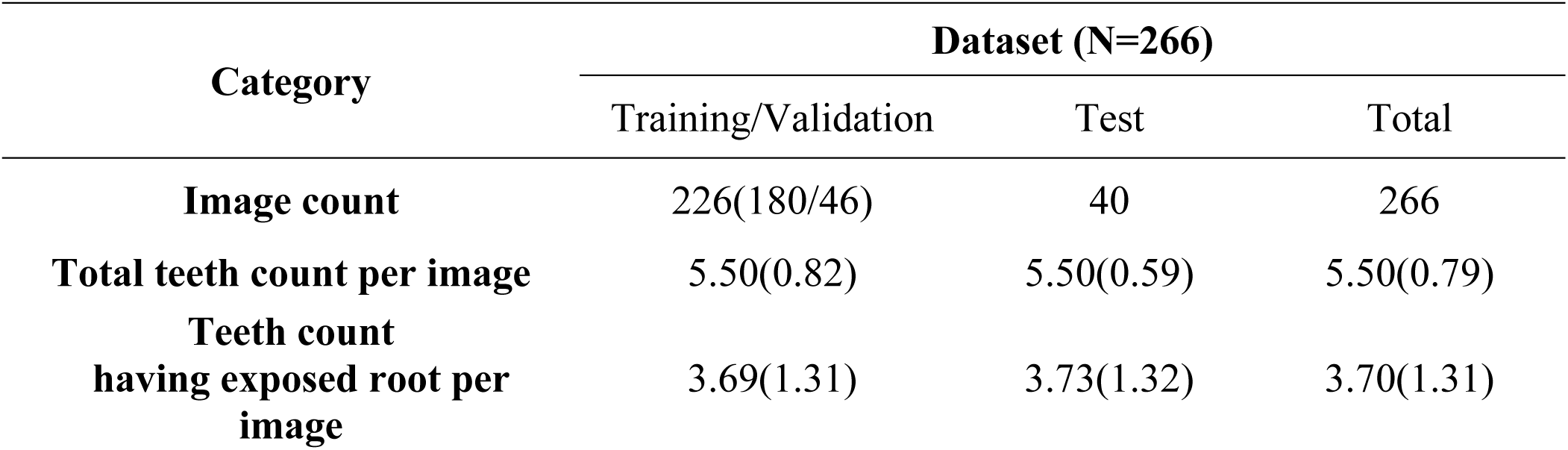
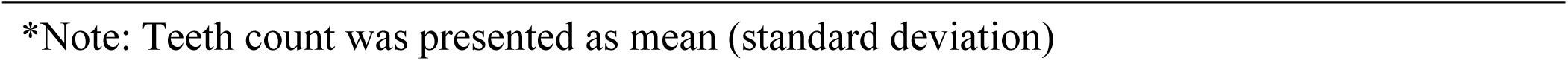
Dataset characteristics.

| Category | Dataset (N=266) |  |  |
| --- | --- | --- | --- |
|  | Training/Validation | Test | Total |
| **Image count** | 226(180/46) | 40 | 266 |
| **Total teeth count per image** | 5.50(0.82) | 5.50(0.59) | 5.50(0.79) |
| **Teeth count**<br>**having exposed root per**<br>**image** | 3.69(1.31) | 3.73(1.32) | 3.70(1.31) |

\*Note: Teeth count was presented as mean (standard deviation)

### 3.2. Model training

Fig 2 presents the results of 5-fold cross-validation for the U-Net++ and YOLOv11 model. Across folds, performance indices including F1-score, Precision, Recall, IOU, and mAP50 remained relatively stable without major fluctuations. Tooth segmentation consistently achieved high accuracy with minimal variation, while root segmentation showed slightly larger variability but remained within a comparable range across folds. The coefficient of variation (CV) further confirmed this trend, with low CV values for tooth segmentation and moderately higher but consistent values for root segmentation. Overall, the results indicate that the model training across folds demonstrates relatively reproducible performance.

**Fig 2.**
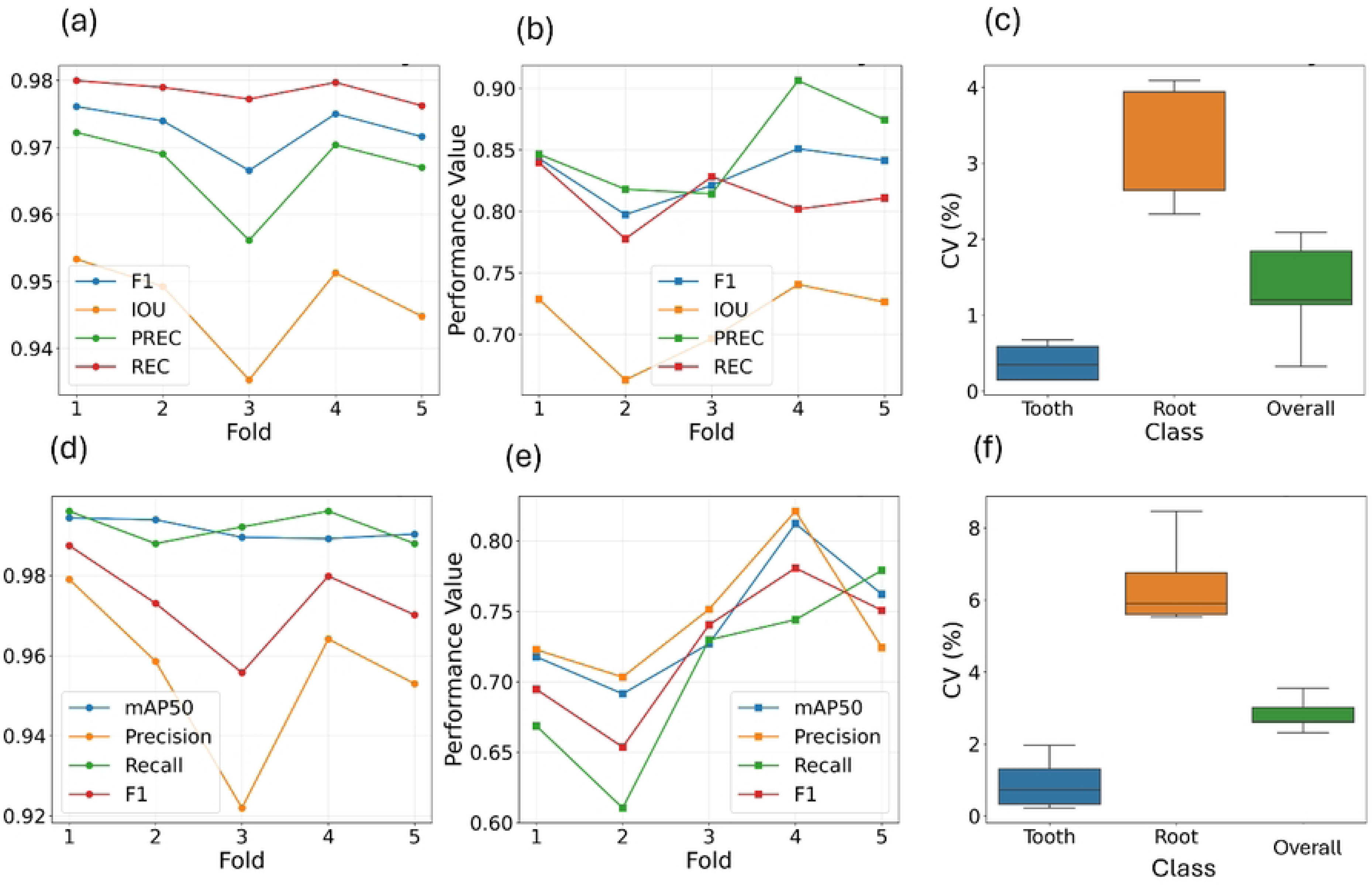
Results of 5-fold cross-validation for the proposed deep learning model in tooth and root segmentation. (a) performance of U-Net++ of tooth segmentation, (b) performance of U-Net++ of root segmentation, (c) coefficient of variation of U-Net++ according to class, (d) performance of YOLO of tooth segmentation, (e) performance of YOLO of root segmentation, (f) coefficient of variation of YOLO according to class

### 3.3. Comparison of Model performance

Performance metrics of YOLO and U-Net++ models on the test dataset are summarized in Table 2. For tooth segmentation, U-Net++ demonstrated superior performance across most indicators, with significantly higher accuracy (0.981 vs. 0.916), Dice score (0.971 vs. 0.936), IoU (0.944 vs. 0.882), and specificity (0.981 vs. 0.968). Both models achieved near-perfect mAP50 values (1.000 for U-Net++ and 0.999 for YOLO), while precision and recall showed minimal differences.

**Table 2.**
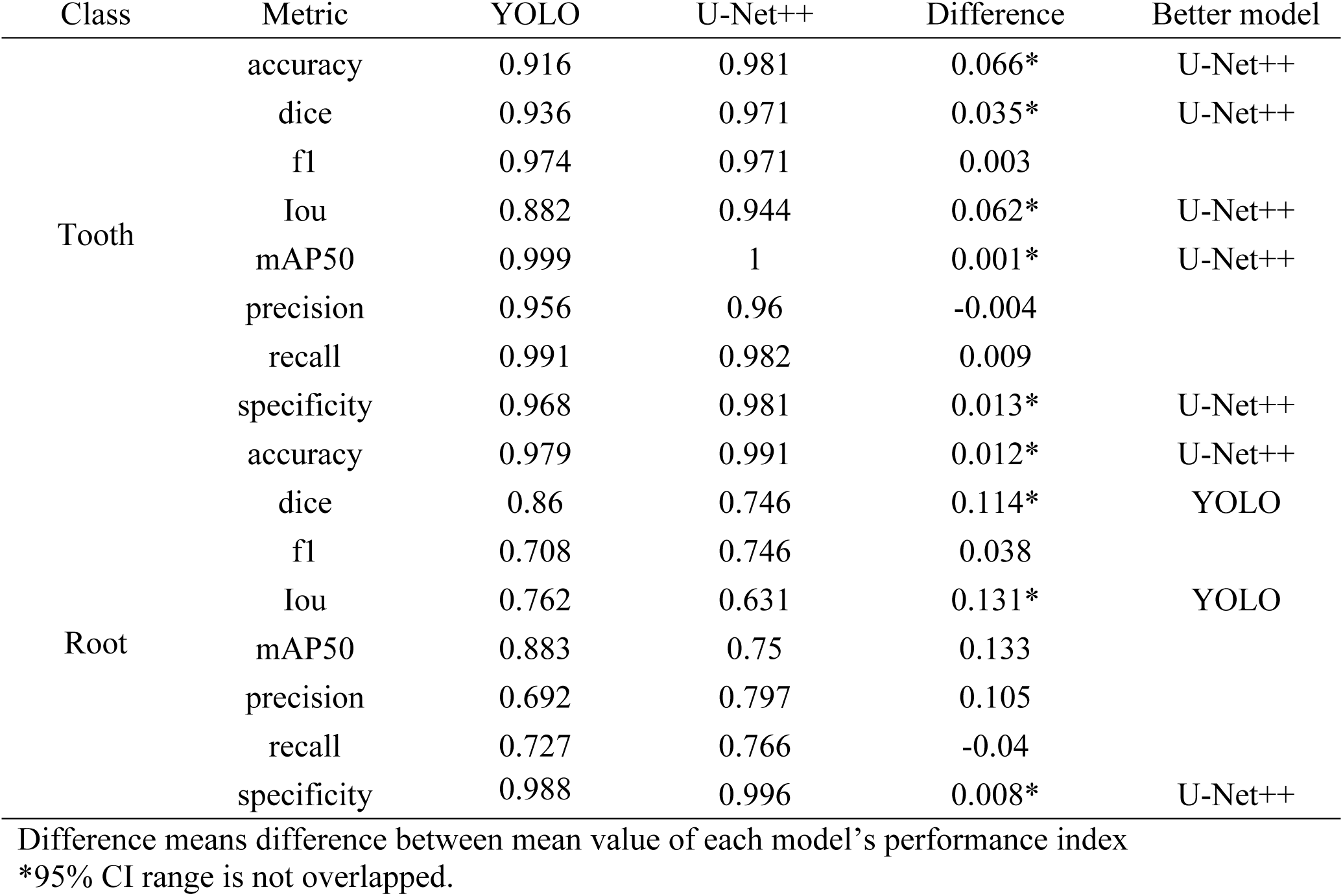
Compare of performance metrics using test dataset according to tooth and root.

In contrast, for root segmentation, YOLO outperformed U-Net++ in terms of Dice score (0.860 vs. 0.746) and IoU (0.762 vs. 0.631), suggesting better localization of root regions. However, U-Net++ achieved slightly higher accuracy (0.991 vs. 0.979) and specificity (0.996 vs. 0.988). Precision and recall were variable across models, with YOLO showing lower precision (0.692) compared to U-Net++ (0.797), but comparable recall (0.727 vs. 0.766).

Taken together, U-Net++ exhibited stronger performance for tooth segmentation, whereas YOLO showed advantages in root segmentation, particularly for overlap-based metrics (Dice and IoU). The qualitative results of the test dataset are presented in Fig 3, which demonstrates the segmentation performance of each model in representative cases.

**Fig 3.**
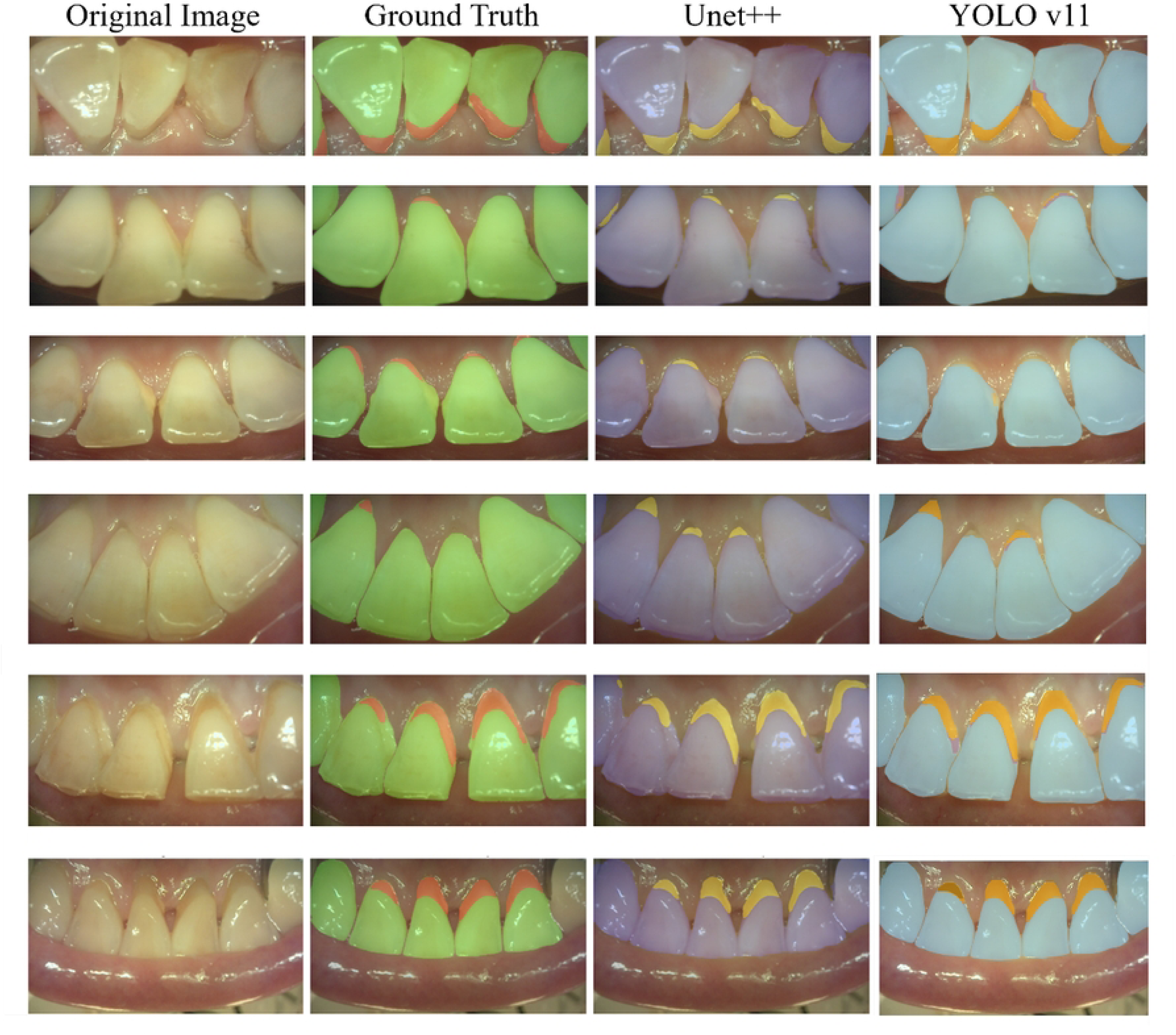
Test visualization of original image and corresponding tooth and root mask according to ground truth and each prediction of U-Net++, and YOLO.

### 3.4. Agreement of quantification of root-to-tooth area

Both U-Net++ and YOLO showed excellent agreement with the ground truth across all metrics (Table 3), with YOLO consistently outperforming U-Net++. For the ERR, tooth area, and root area, YOLO achieved higher CCC (0.973–0.978) and ICC (0.975–0.978) values compared to U-Net++ (CCC: 0.933–0.968, ICC: 0.927–0.960). These findings indicate that while both models are reliable, YOLO provides more superior quantification agreement to ground truth.

**Table 3.** Agreement of tooth, exposed root ratio (ERR) between ground truth and each model’s prediction.

| Category | Model | CCC |  | ICC |  |
| --- | --- | --- | --- | --- | --- |
|  |  | Mean<br>n | [95%CI] | Mean | [95%CI] |
| Exposed Root Ratio (ERR) | U-Net++ | 0.933 | [0.887, 0.976] | 0.927 | [0.926, 0.928] |
|  | YOLO | 0.973 | [0.951, 0.984] | 0.975 | [0.974, 0.976] |
| Tooth Area | U-Net++ | 0.968 | [0.944, 0.985] | 0.96 | [0.959, 0.961] |
|  | YOLO | 0.978 | [0.964, 0.990] | 0.976 | [0.975, 0.977] |
| Root Area | U-Net++ | 0.953 | [0.917, 0.983] | 0.953 | [0.952, 0.954] |
|  | YOLO | 0.977 | [0.956, 0.988] | 0.978 | [0.977, 0.979] |

\*Note: CCC means concordance correlation coefficient and ICC means intraclass correlation coefficient

## 4. Discussion

This study developed and compared two leading deep learning models, YOLOv11 and U-Net++, for segmenting teeth and exposed roots in intraoral camera images of the mandibular anterior lingual region, utilizing a dataset of 266 images from 291 patients with a mean age of 52.8 years. Considering relatively small number of the dataset, the 5-fold cross-validation was applied and their results demonstrated stable performance with low coefficients of variation for tooth segmentation and moderate variability for exposed roots, confirming reproducibility. Also, both models exhibited favored performance consistently despite data variability, enhanced by data augmentation and hyperparameter optimization. In particular, in case of exposed root quantification, a ratio of exposed root area to tooth area, high agreement between AI models and ground truth was confirmed, which suggest their clinical and academic applicability.

Specifically, superiority of U-Net++ in tooth segmentation and YOLOv11 in root segmentation was observed respectively, throughout direct comparison of pixel-based and object-based AI models. YOLOv11’s higher performance for detection of root exposure might contribute to higher agreement in exposed root quantification compared to U-Net++. This difference is likely attributable to the distinct architectural characteristics of the models. U-Net++, with its encoder-decoder structure and dense skip connections, is well suited for preserving fine spatial information and segmenting relatively large and continuous anatomical structures such as tooth crowns and roots as a whole. This is consistent with previous studies reporting strong performance of U-Net-based models for tooth segmentation in dental imaging [29,41,42]. In contrast, YOLOv11 performs instance segmentation and is optimized for localizing small, irregularly shaped, and visually heterogeneous targets [25]. This may explain its superior performance in detecting exposed root areas, which are often limited in extent and affected by anatomical variation, plaque, calculus, or image-quality differences. The higher Dice coefficient and IoU for root segmentation observed with YOLOv11 support this interpretation.

In terms of quantitative agreement, YOLOv11 showed stronger concordance with the ground truth than U-Net++ for the ERR (CCC, 0.973 vs. 0.933; ICC, 0.975 vs. 0.927), based on bootstrap analysis. These findings suggest that accurate localization of exposed roots is critical for reliable quantification and that YOLOv11 may be more suitable when the primary goal is objective measurement rather than broad semantic segmentation. At the same time, the lower precision of YOLOv11 for roots relative to U-Net++ indicates that some over-detection may occur, which is a common trade-off in detection-oriented models. Conversely, the lower root performance of U-Net++ may reflect the difficulty of learning sparse and visually subtle root exposure regions in a pixel-wise framework, especially under class imbalance.

From a clinical perspective, quantification of the ERR may provide an objective and non-invasive imaging biomarker related to gingival recession and periodontal tissue loss [22]. Because lingual root exposure in the mandibular anterior region has been associated with chronic periodontitis, this metric may be useful for screening, longitudinal monitoring, and patient education. AI-assisted quantification of root exposure from intraoral camera images may serve as a useful adjunct to conventional periodontal examination, especially in settings where radiographic evaluation is not readily available or where repeated non-invasive monitoring is preferred. However, it should not be viewed as a substitute for comprehensive periodontal assessment, which requires clinical examination and consideration of systemic and behavioral risk factors such as smoking, diabetes, and oral hygiene status [43].

This study has several limitations. First, the dataset was derived from a single institution and the test set was relatively small, which may limit generalizability. Second, image quality may have been influenced by lighting conditions, debris, calculus, saliva, and variation in acquisition angle, all of which can affect segmentation accuracy. Third, the study focused on the mandibular anterior lingual region and did not include broader anatomical regions or sufficiently diverse disease severities. Nevertheless, the use of repeated bootstrap resampling strengthened the reliability of the performance estimates despite the limited sample size.

Future studies should include larger, multi-center datasets with greater demographic and clinical diversity to validate the generalizability of these findings. In addition, inclusion of mild or early-stage cases, standardized image acquisition protocols, and prospective clinical validation would further clarify the role of AI-based root exposure quantification in periodontal screening and monitoring.

## 5. Conclusions

This study evaluated YOLOv11 and U-Net++ for quantifying tooth and root exposure on intraoral camera images of the mandibular anterior lingual region. U-Net++ showed superior performance for tooth segmentation, whereas YOLOv11 achieved better performance for root segmentation, with higher Dice coefficient (0.860) and IoU (0.762). YOLOv11 also demonstrated stronger agreement with the ground truth for quantifying the ERR (CCC, 0.973; ICC, 0.975), indicating that it is a promising model for root exposure quantification in this setting.

Overall, these findings suggest that YOLOv11-based analysis of intraoral images may support objective assessment of exposed root areas and may have potential as an adjunctive tool for periodontal screening and monitoring. Further validation using larger and more diverse datasets is needed before routine clinical application.

## Data Availability

Due to ethical and privacy restrictions, the raw dataset (intraoral images) is not publicly available. Data may be requested from the corresponding author upon reasonable request, subject to approval by the Institutional Review Board of Yeungnam University Hospital.

## Ethical Approval

The study was conducted in accordance with the Declaration of Helsinki and approved by the Institutional Review Board (IRB) of Yeungnam University Hospital (YUMC IRB 2021-07-019-002).

## Informed Consent

Informed consent was obtained from all subjects involved in the study and written informed consent has been obtained from the patients to publish this paper.

